# Automated, Objective Speech and Language Markers of Longitudinal Changes in Psychosis Symptoms

**DOI:** 10.1101/2024.07.19.24310718

**Authors:** Sunny X. Tang, Michael J. Spilka, Majnu John, Michael L. Birnbaum, Ema Saito, Sarah A. Berretta, Leily M. Behbehani, Mark Y. Liberman, Anil K. Malhotra, William Simpson, John M. Kane

**Author notes:** Corresponding Author: Sunny X. Tang, M.D. Zucker Hillside Hospital 75-59 263^rd^ St., Glen Oaks, NY 11004.

## Abstract

**Background and Hypotheses:** We sought to evaluate the ability of automated speech and language features to track fluctuations in the major psychosis symptoms domains: *Thought Disorder, Negative Symptoms*, and *Positive Symptoms*.

**Study Design:** Sixty-six participants with psychotic disorders were longitudinally assessed soon after inpatient admission, at discharge, and at 3- and 6-months. Psychosis symptoms were measured with semi-structured interviews and standardized scales. Recordings were collected from paragraph reading, fluency, picture description, and open-ended tasks. Longitudinal relationships between psychosis symptoms and 357 automated speech and language features were analyzed using a single component score and as individual features, using linear mixed models.

**Study Results:** All three psychosis symptom domains demonstrated significant longitudinal relationships with the single component score. *Thought Disorder* was particularly related to features describing more subordinated constructions, less efficient identification of picture elements, and decreased semantic distance between sentences. *Negative Symptoms* was related to features describing decreased speech complexity. *Positive Symptoms* appeared heterogeneous, with *Suspiciousness* relating to greater use of nouns, and *Hallucinations* related to decreased semantic distances. These relationships were largely robust to interactions with gender and race. However, interactions with timepoint revealed variable relationships during different phases of illness (acute vs. stable).

**Conclusions:** Automated speech and language features show promise as scalable, objective markers of psychosis severity. The three symptom domains appear to be distinguishable with different features. Detailed attention to clinical setting and patient population is needed to optimize clinical translation; there are substantial implications for facilitating differential diagnosis, improving psychosis outcomes and enhancing therapeutic discovery.

## Introduction

Psychotic disorders are severe mental illnesses and include schizophrenia spectrum disorders as well as bipolar and major depressive disorders with psychotic features; the total lifetime prevalence is 2-3% (1). While psychotic disorders are associated with significant disability, increased health care costs, family burden, and reduced life expectancy in general (2), outcomes are heterogenous and can be improved with a range of effective treatments. Antipsychotic medications remain a mainstay of pharmacologic treatment and are effective against a range of psychosis symptoms, but benefits can be limited by non-response, non-adherence, and significant side effects (3). Psychosocial treatments like cognitive remediation, social skills training, psychotherapy, and self-management, as well as multi-disciplinary early-intervention programs have also demonstrated efficacy (4,5). There is a great deal of interest in developing approaches to ‘precision psychiatry’, whereby objective biomarkers can be used to stratify patients, optimize treatment decisions and provide patients with more effective and timely care (6).

Natural language processing (NLP) and speech and language features evaluated with automated, computerized methods may offer substantial advantages as a scalable, cost-effective, low-burden means for generating clinically-relevant markers for psychosis. These methods generate a range of objective features describing the timing (e.g., latency, speaking rate), acoustic properties (e.g., frequency, amplitude), lexical characteristics (e.g., sentiment, commonness), and structure (e.g., syntax, semantic coherence, speech graph properties) of speech. They require relatively little expertise or specialized equipment to capture and, when fully developed, can be implemented in a cost- and time-efficient manner, relying on automated computer algorithms (7). There is now consistent evidence demonstrating that a range of objective speech and language features can be used as objective markers of psychosis. These methods are highly sensitive (8) and consistently predict schizophrenia diagnosis relative to healthy controls, as well as conversion to psychosis among individuals at clinical high risk (9). Different types of speech and language features are also sensitive to different dimensions of psychosis symptoms, cognition, and functioning (10–13). One study used a range of lexical, coherence, and disfluency features to longitudinally estimate psychosis symptoms in 38 participants with psychotic disorders (99 total sessions) and found promising between- and within-participant relationships to positive and negative symptoms (14). However, in general, there are few studies evaluating the longitudinal relationships between fluctuations in psychosis symptoms and objective speech and language features, knowledge of which is necessary to support the development of speech-based clinical monitoring applications.

Here, we sought to evaluate the ability of automated speech and language features to track fluctuations in psychosis symptoms among 66 participants with psychotic disorders over 160 sessions (up to 4 timepoints per participant). The long-term goal of this work is to develop a means for measuring “vital signs” in psychosis – i.e., sensitive, objective measures of psychosis severity which can be obtained rapidly and cost-effectively. We apply a broad approach, integrating information from a wide range of speech and language features assessed via several task contexts. The clinical outcomes of interest were the principal psychosis symptom domains: 1) *Thought Disorder / Disorganization*, which are early signs of both relapse and treatment response for psychosis (15,16) and can be directly related to speech and language disturbance; 2) *Negative Symptoms*, which has significant implications for functional outcomes (2) and also includes speech-related constructs; and 3) *Positive Symptoms*, which are important targets of antipsychotic treatment and predictors of rehospitalization (17). We first evaluate speech and language features in general, as a single component score, and then examine relationships with individual speech and language features.

## Methods

### Participants

Recruitment occurred on acute inpatient psychiatric units at the Zucker Hillside Hospital in Glen Oaks, NY. Inclusion criteria were age 15-40 years, proficient in English, current diagnosis of bipolar I disorder with psychotic features or schizophrenia spectrum disorder (schizophrenia, schizophreniform disorder, schizoaffective disorder, unspecified psychotic disorder, or brief psychotic disorder), onset of illness within 2 years, and at least moderate positive or disorganized symptoms on admission. Young patients at early phases of illnesses were selected due to greater anticipated fluctuations in severity. Individuals with substance-induced psychotic disorders were excluded, along with those with comorbidities directly affecting speech production or language ability (e.g., aphasia, stroke, autism spectrum disorder). The research procedures were approved by the institutional review board at Northwell Health, and all participants provided written consent after decisional capacity was confirmed. The study was registered on ClinicalTrials.gov (NCT05601050).

Two participant sessions were impacted by poor recording environment and therefore excluded from the analyses. A total of 66 participants and 160 sessions are described here (Table 1).

**Table 1.** Participant Characteristics.

|  | **Baseline** | **Discharge** | **3mo** | **6mo** | **p value** |
| --- | --- | --- | --- | --- | --- |
| **n** | 66 | 54 | 22 | 18 |  |
| **Age (SD)** | 26.4 (5.3) | 26.1 (4.7) | 27.0 (6.0) | 27.4 (5.2) | 0.77 |
| **Sex (%)** |  |  |  |  | 0.31 |
| Female | 20 (30%) | 15 (28%) | 2 (9%) | 2 (11%) |  |
| Intersex | 1 (2%) | 1 (2%) | 0 (0%) | 0 (0%) |  |
| Male | 45 (68%) | 38 (70%) | 20 (91%) | 16 (89%) |  |
| **Gender (%)** |  |  |  |  | 0.36 |
| Man | 45 (75%) | 38 (78%) | 19 (91%) | 15 (88%) |  |
| Woman | 15 (25%) | 11 (22%) | 2 (10%) | 2 (12%) |  |
| Not Reported | 6 | 5 | 1 | 1 |  |
| **Race (%)** |  |  |  |  | 0.48 |
| Asian | 12 (18%) | 11 (21%) | 4 (18%) | 3 (17%) |  |
| Black/African American | 28 (43%) | 24 (45%) | 11 (50%) | 12 (67%) |  |
| Other | 9 (14%) | 7 (13%) | 0 (0%) | 0 (0%) |  |
| White/Caucasian | 16 (25%) | 11 (21%) | 7 (32%) | 3 (17%) |  |
| Not Reported | 1 | 1 | 0 | 0 |  |
| **Ethnicity** |  |  |  |  | 0.60 |
| Hispanic | 10 (15%) | 6 (11%) | 1 (5%) | 0 (0%) |  |
| Not Hispanic | 52 (79%) | 45 (83%) | 20 (91%) | 17 (94%) |  |
| Not Reported | 4 | 3 | 1 | 1 |  |
| **Education (SD)** | 14.1 (1.9) | 14.0 (1.8) | 14.1 (1.8) | 14.0 (1.7) | 0.99 |
| **Diagnosis** |  |  |  |  | 0.99 |
| Bipolar w. Psychosis | 4 (6%) | 3 (6%) | 1 (5%) | 1 (6%) |  |
| Schizoaffective | 15 (23%) | 13 (24%) | 4 (18%) | 3 (17%) |  |
| Schizophrenia | 33 (50%) | 29 (54%) | 11 (50%) | 11 (61%) |  |
| Schizophreniform | 4 (6%) | 2 (4%) | 2 (9%) | 0 (0%) |  |
| Unspecified PD | 10 (15%) | 7 (13%) | 4 (18%) | 3 (17%) |  |
| **TLC Total (SD)** | 22.0 (14.4) | 15.6 (11.3) | 11.7 (12.0) | 12.7 (11.4) | 0.001 |
| **SANS Total Global (SD)** | 8.3 (3.5) | 7.2 (3.4) | 7.5 (3.8) | 7.1 (4.9) | 0.47 |
| **BPRS Positive Symptoms (SD)** | 20.7 (5.4) | 16.7 (6.4) | 14.5 (6.6) | 13.5 (6.8) | <0.001 |

Note: BPRS – Brief Psychiatric Rating Scale; SANS – Scale for the Assessment of Negative Symptoms; SD – standard deviation; TLC – Scale for the Assessment of Thought Language and Communication

### Assessments

Participants were assessed longitudinally over 4 sessions. The first session (baseline) was conducted as soon as possible after participants were admitted. The second session (discharge) was conducted when imminent discharge was planned or within 1 week after discharge. To limit variability, a range of 1 – 3 weeks was imposed for the interval between the first and second sessions, reflecting average hospitalization durations and when the greatest clinical change is expected. The third and fourth sessions were conducted at 3 months and 6 months after discharge.

Diagnoses were confirmed with the SCID-IV-TR (18) using DSM-5 criteria. *Thought Disorder* was rated with the Scale for the Assessment of Thought, Language and Communication (TLC), and total score was calculated (19). *Negative Symptoms* were rated with the Scale for the Assessment of Negative Symptoms (SANS), and global scores were totaled for the Affective Flattening, Alogia, Avolition/Apathy, and Anhedonia/Asociality domains per Robinson et al. (20,21). *Positive Symptoms* were rated with the Brief Psychiatric Rating Scale (BPRS) and the factor score was calculated per Overall et al. (22) totaling these items: Hostility, Suspiciousness, Uncooperativeness, Hallucinatory Behavior, Conceptual Disorganization, and Unusual Thought Content. All clinical assessments were conducted by trained assessors who underwent departmental training to establish reliability.

Speech was collected using iPads with the Winterlight iOS application. Participants were asked to respond to 4 tasks, sometimes using multiple stimuli (Figure 1A): paragraph reading (standardized text), fluency (animal category fluency, F-letter phonemic fluency), picture descriptions (3 pictures per session), and open-ended journaling (2 self-descriptive narrative prompts). The full assessment took 10-15 minutes to complete. Responses were audio-recorded, transcribed with a combination of automated processes and human annotation, and processed through an automated pipeline for extracting speech and language features using the Winterlight platform (winterlightlabs.com).

**Figure 1.**
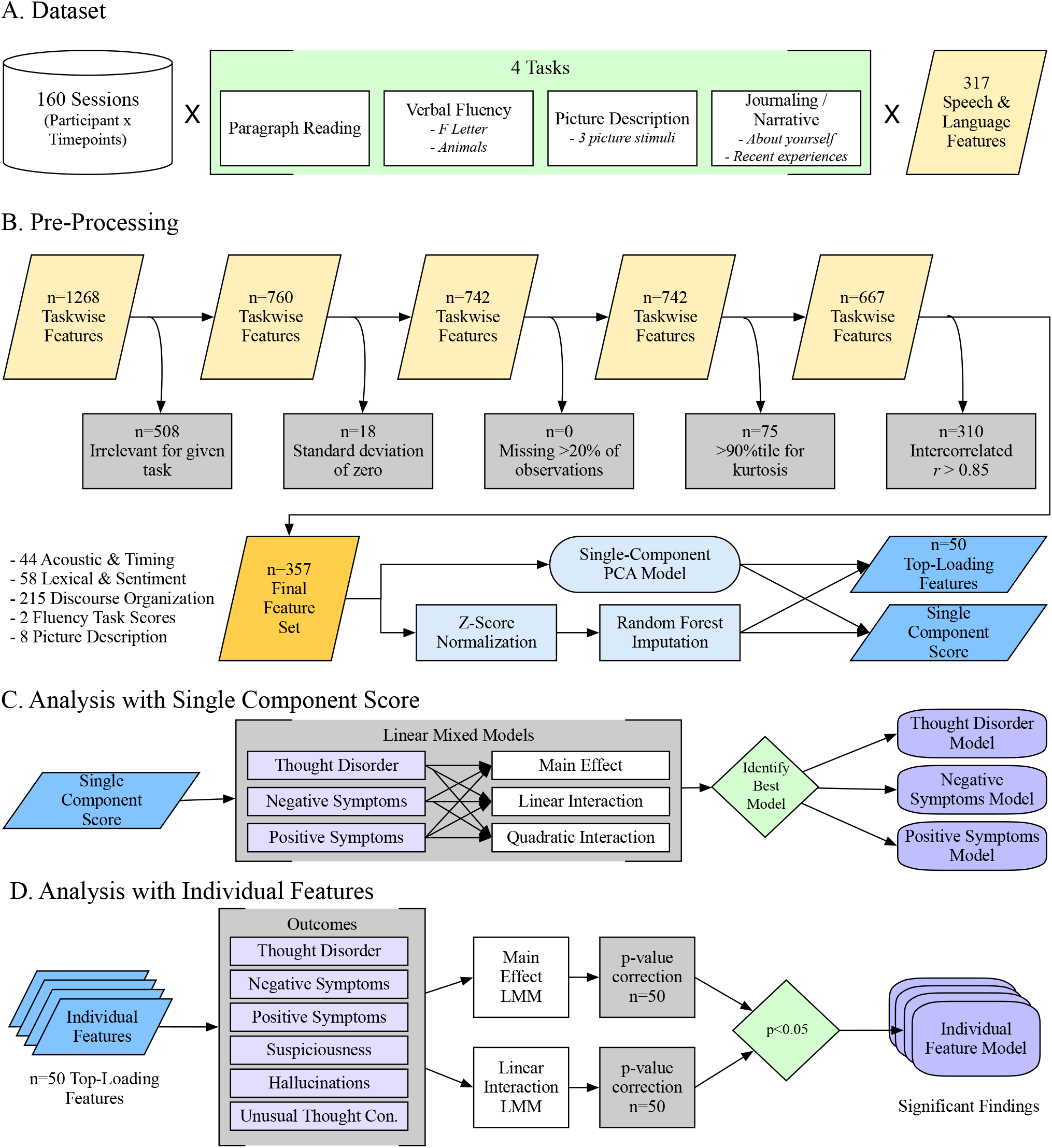
Flowchart for Data Processing and Analyses. Note: PCA – Principal Component Analysis; LMM – Linear mixed model.

### Speech and Language Features

The automated pipeline produced 317 raw speech and language features for each stimulus (Figure 1A; 45 acoustic and timing, 27 lexical characteristics, 216 discourse organization, 2 fluency task scores, 27 picture description content measures). Features were collapsed to the task-level (i.e., each task within each session was evaluated separately); where there were multiple stimuli for one task (e.g., 3 different pictures), the features were averaged across the stimuli. This produced 1,268 task-wise features (Figure 1B). A series of features were then excluded to remove those that were not task-relevant (e.g., syntactic features removed for paragraph reading and fluency tasks), lacked sufficient variability (based on standard deviation and kurtosis), or too highly intercorrelated (a Spearman correlation matrix was calculated, features were ranked from highest to lowest by total correlations with all other features, then successively excluded if they were correlated *r*>0.85 with another feature). The final feature set included 357 task-wise features (44 acoustic and timing, 28 lexical characteristics, 215 discourse organization, 2 fluency task scores, 8 picture description content measures).

A single component score was calculated to represent the speech and language features globally. This process involved: First, we performed a principal component analysis (PCA) on unimputed data using pairwise deletions (12 features missing up to 6 observations each), resulting in a 1-component model explaining 6.7% of the total variance. Then, we imputed missing data using random forest imputation with *missForest* v.1.5 in R. Finally, we extracted a single component score for each participant observation, represented as a z-score. The PCA and component score extraction was completed using *psych* v.2.2.5 in R.

Subsequently, we examined relationships between psychosis symptom domains and individual speech features. Out of consideration for multiple comparisons, the 50 top-loading features from the PCA (described in Supplemental Table 1) were selected as candidates because they were most representative of the single component score. However, selecting candidate features in this way biases toward over-representation of tasks and feature types that were more common among the final feature set, i.e., picture description and journaling tasks, and discourse organization features.

### Statistical Analyses

To understand how psychosis is longitudinally related to speech and language features globally, each psychosis symptoms domain (*Thought Disorder, Negative Symptoms, Positive Symptoms*) was predicted using random-intercept linear mixed models (LMMs) with the single component score and timepoint as fixed effects, and participant as the observation unit for random effects (Figure 1C). The main effect of the component score and linear and quadratic interactions with timepoint were examined (Supplemental Table 2 details model structures). The default unstructured variance-covariance structure within nlme R package was used in all LMM analyses (23). Timepoint was centered around the baseline, and model fit is reported with the Akaike and Bayesian Information Criteria (AIC and BIC), with greater emphasis on the BIC because it incorporates the sample size also into the penalty term.

Each psychosis domain was then predicted with LMMs for the 50 top-loading individual features to better understand the contribution of specific speech and language features (Figure 1D). Quadratic interaction models were not examined because they were not significant for any of the single component LMMs. To account for multiple comparisons with 50 features, p-values for the parameter of interest were adjusted using the Benjamini and Hochberg false-discovery rate (FDR) method (24). Because there were no findings for the *Positive Symptoms* domain that survived FDR correction, we hypothesized that *Positive Symptom*s may be too heterogeneous as a clinical construct and further tested individual symptoms. *Hallucinations, Suspiciousness*, and *Unusual Thought Content* were evaluated because these demonstrated sufficient variance in the sample and represent core positive symptoms.

Interactions with race and gender were explored by testing the interaction between the parameter of interest and the demographic variables. We did not examine interactions with age because the age range was relatively narrow.

All analyses were conducted in RStudio with R v.4.2.0.

## Results

### Trajectories of Psychosis Symptoms

Changes in psychosis symptoms followed expected patterns (Table 1), with significant declines in *Thought Disorder* (p=0.001) and *Positive Symptoms* (p<0.001). There was no overall effect of timepoint on *Negative Symptoms* (p=0.47). However, there was significant individual variability, as can be observed from the individual datapoints plotted in Figure 2.

**Figure 2.**
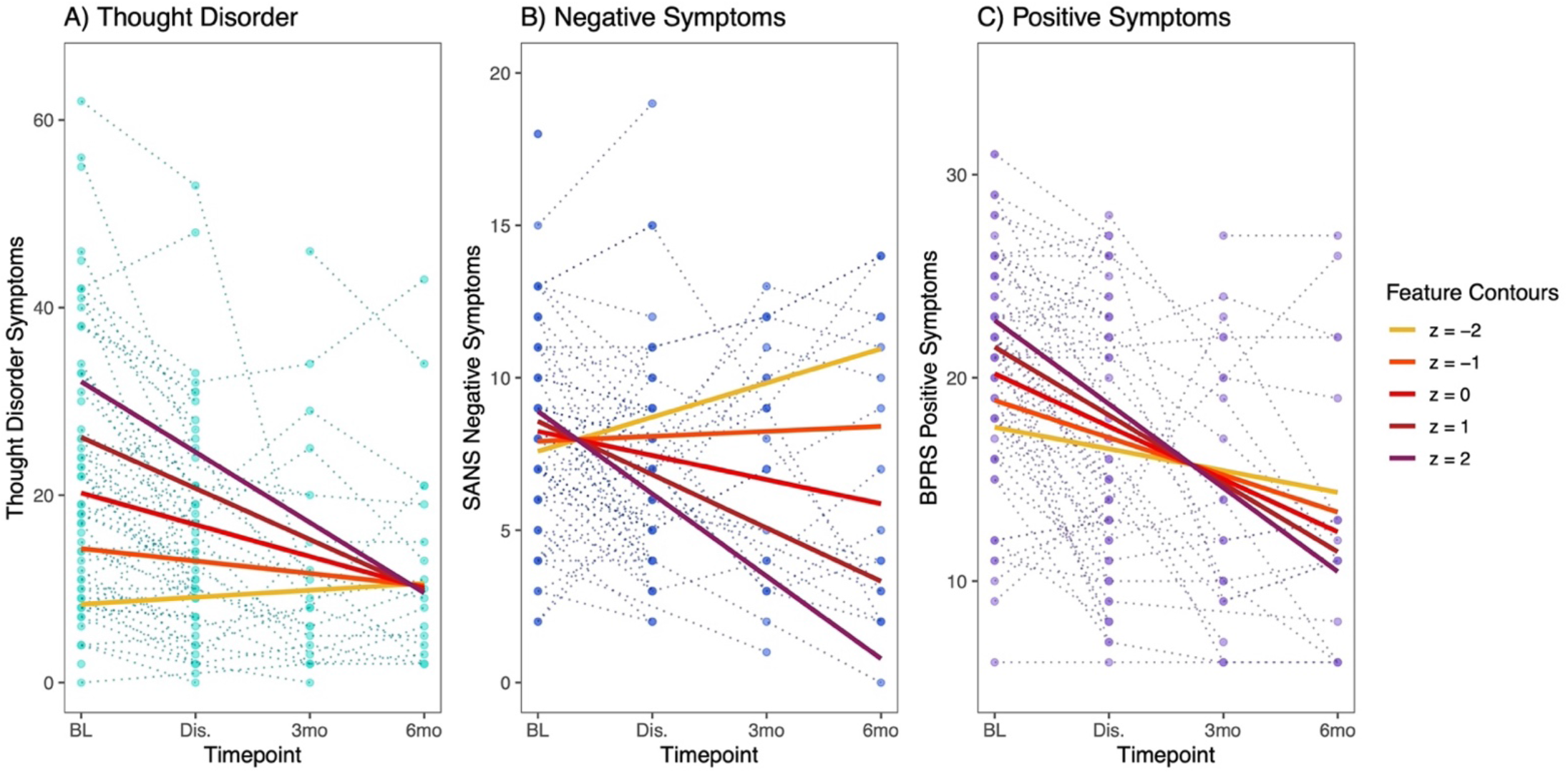
Psychosis Symptoms and Single Component Score. The single component score (“Feature”) represents the overall variance from the 357 speech and language measures in the final feature set. Variance predicted by the single component score in LMMs is shown for A) *Thought Disorder* – total TLC score, B) *Negative Symptoms* – total global SANS scores, and C) *Positive Symptoms* – BPRS factor score. For all three symptom domains, the best fit model was the LMM including a linear interaction term between the single component score and timepoint. In each subplot, individual observations for each participant are plotted across the 4 timepoints (*Thought Disorder* – turquoise, *Negative Symptoms* – blue, *Positive Symptoms* – purple). Feature contours illustrate LMM predictions for each symptom domain at different values of the single component score (z=-2 to +2) across the 4 timepoints. When contour lines are farther apart, greater variance in symptom severity is predicted by the single component score. For example, at baseline, total TLC score (*Thought Disorder)* is estimated by the LMM to be ∼30 for individuals with single component score z=2, while total TLC score is estimated at ∼10 for individuals with single component score z=-2. At 6mo follow-up, there is very little difference in the estimated *Thought Disorder* severity regardless of the single component score.

### Psychosis Domains and Single Component Score

There were significant longitudinal relationships between the single component score and all three psychosis symptoms domains (Table 2; unabbreviated results - Supplemental Table 3). For *Thought Disorder*, the main effect for the single component score was significant in all three models, in addition to the linear interaction term. The linear interaction model was chosen as the best fit based on the BIC. For *Negative Symptoms* and *Positive Symptoms*, only the linear interaction term was significant. The relationships between the single component score and psychosis symptoms are illustrated in Figure 2. Significant individual variability can be observed for all symptom domains, with feature contours indicating the predicted symptom severity for different values of the single component score, across timepoints. Note that for *Thought Disorder*, the greatest variance is predicted from the component score at baseline and discharge, compared to *Negative Symptoms* where the greatest variance is predicted at 3- and 6-month follow-up. Higher component scores are related to greater *Thought Disorder* but less *Negative Symptoms*. For *Positive Symptoms*, the polarity of the relationship reverses after discharge.

**Table 2.** Linear Mixed Models Results.

**A. Features Relating to Thought Disorder**
|  |  | Int. | Timept. | Main Effect |  |  | Lin. Interaction |  |  | Model |  |
| --- | --- | --- | --- | --- | --- | --- | --- | --- | --- | --- | --- |
| Feature | Model | Coeff. | Coeff. | Coeff. | p | p-adj. | Coeff. | p | p-adj. | AIC | BIC |
| Single component score | L | 20.2 | -3.4 | 5.9 | <0.001 |  | -2.1 | <0.001 |  | 1212 | 1230 |
| Subordinating conjunctions (JOU) | M | 20.6 | -3.3 | 4.2 | <0.001 | <0.001 |  |  |  | 1212 | 1227 |
| Min. utterance semantic dist. (Google) (JOU) | M | 20.8 | -3.5 | -3.5 | <0.001 | 0.001 |  |  |  | 1216 | 1231 |
| Min. utterance semantic distance (fastText) (PIC) | M | 20.9 | -3.7 | -3.3 | <0.001 | 0.002 |  |  |  | 1218 | 1233 |
| Picture units identified:Action (PIC) | M | 21.6 | -4.3 | -3.6 | <0.001 | 0.002 |  |  |  | 1218 | 1234 |
| Total audio duration (JOU) | M | 20.8 | -3.5 | 3.5 | <0.001 | 0.002 |  |  |  | 1219 | 1234 |
| Picture units identified (PIC) | L | 21.0 | -3.8 | -5.6 | <0.001 |  | 2.2 | <0.001 | 0.03 | 1214 | 1232 |
| Noun phrase: Noun (JOU) | L | 21.1 | -4 | -4.3 | <0.001 |  | 2.3 | 0.002 | 0.03 | 1219 | 1238 |

**B. Features Relating to Negative Symptoms**
|  |  | Int. | Timept. | Main Effect |  |  | Lin. Interaction |  |  | Model |  |
| --- | --- | --- | --- | --- | --- | --- | --- | --- | --- | --- | --- |
| Feature | Model | Coeff. | Coeff. | Coeff. | p | p-adj. | Coeff. | p | p-adj. | AIC | BIC |
| Single component score | L | 8.2 | -0.8 | 0.3 | 0.38 |  | -1.0 | <0.001 |  | 844 | 862 |
| Subordinate clause: preposition + sentence (JOU) | L | 8.2 | -0.6 | 1.1 | 0.003 |  | -0.9 | <0.001 | 0.02 | 850 | 868 |
| Min. utterance semantic distance (fastText) (PIC) | L | 8.2 | -0.6 | -0.4 | 0.2 |  | 0.7 | 0.001 | 0.02 | 851 | 870 |
| Imageability (PIC) | L | 8.3 | -0.5 | -0.5 | 0.2 |  | 0.7 | 0.002 | 0.02 | 852 | 870 |
| Age of acquisition (PIC) | L | 8.1 | -0.4 | 0.6 | 0.1 |  | -0.7 | 0.002 | 0.02 | 853 | 871 |
| Adjective phrase length (PIC) | L | 8.3 | -0.7 | 0.1 | 0.7 |  | -0.9 | 0.003 | 0.02 | 847 | 865 |
| Average utterance semantic distance (Google) (PIC) | L | 8.2 | -0.6 | -0.1 | 0.8 |  | 0.6 | 0.003 | 0.02 | 850 | 868 |
| Number of edges (JOU) | L | 8.1 | -0.5 | -0.7 | 0.05 |  | 0.9 | 0.004 | 0.02 | 854 | 872 |

**C. Features Relating to Positive Symptoms**
|  |  | *Int.* | *Timept.* | *Main Effect* |  |  | *Lin. Interaction* |  |  | *Model* |  |
| --- | --- | --- | --- | --- | --- | --- | --- | --- | --- | --- | --- |
| *Feature* | *Model* | *Coeff.* | *Coeff.* | *Coeff.* | *p* | *p-adj.* | *Coeff.* | *p* | *p-adj.* | *AIC* | *BIC* |
| Single component score | L | 20.2 | -2.6 | 1.3 | 0.03 |  | -0.8 | 0.04 |  | 997 | 1015 |

**D. Features Relating to Suspiciousness**
|  |  | *Int.* | *Timept.* | *Main Effect* |  |  | *Lin. Interaction* |  |  | *Model* |  |
| --- | --- | --- | --- | --- | --- | --- | --- | --- | --- | --- | --- |
| *Feature* | *Model* | *Coeff.* | *Coeff.* | *Coeff.* | *p* | *p-adj.* | *Coeff.* | *p* | *p-adj.* | *AIC* | *BIC* |
| Subordinating conjunctions (JOU) | M | 3.9 | -0.5 | 0.5 | 0.001 | 0.03 |  |  |  | 663 | 678 |
| Noun phrase rate (JOU) | M | 3.9 | -0.5 | -0.5 | 0.002 | 0.03 |  |  |  | 663 | 678 |
| Noun phrase: Noun (JOU) | M | 3.9 | -0.5 | -0.5 | 0.002 | 0.03 |  |  |  | 663 | 678 |
| Imageability (JOU) | M | 3.9 | -0.4 | -0.5 | 0.003 | 0.04 |  |  |  | 664 | 680 |
| Number of edges (JOU) | M | 3.9 | -0.5 | -0.4 | 0.005 | 0.05 |  |  |  | 665 | 680 |

**E. Features Relating to Hallucinations**
|  |  | *Int.* | *Timept.* | *Main Effect* |  |  | *Lin. Interaction* |  |  | *Model* |  |
| --- | --- | --- | --- | --- | --- | --- | --- | --- | --- | --- | --- |
| *Feature* | *Model* | *Coeff.* | *Coeff.* | *Coeff.* | *p* | *p-adj.* | *Coeff.* | *p* | *p-adj.* | *AIC* | *BIC* |
| Age of acquisition: nouns (JOU) | M | 3.6 | -0.5 | -0.5 | 0.001 | 0.06 |  |  |  | 645 | 660 |
| Min. utterance semantic distance (fastText) (PIC) | M | 3.5 | -0.5 | 0.5 | 0.003 | 0.06 |  |  |  | 646 | 662 |
| Average utterance semantic distance (Google) (PIC) | M | 3.5 | -0.4 | 0.5 | 0.003 | 0.06 |  |  |  | 646 | 662 |

*Note:* M – main effect; L – linear effect; JOU – Journaling task; PIC – Picture description task. See Supplemental Table 3 for unabbreviated LMM results, including quadratic interaction models.

### Psychosis Symptoms and Individual Speech and Language Features

Fifty speech and language features were evaluated individually for their longitudinal relationships with the psychosis symptoms, with many significant even after correcting for multiple comparisons (Table 2; unabbreviated results – Supplemental Table 3).

*Thought Disorder* was significantly related to 15 features: 12 through main effects, and 3 through linear interactions with timepoint. These prominently included subordinate sentence constructions with more subordinate clauses reflecting greater *Thought Disorder* (7 related features; e.g., Figure 3A). Lower minimum semantic distance between adjacent sentences (i.e., closer in meaning) and fewer entities correctly identified on picture description tasks (e.g., Figure 3B) were also related to greater *Thought Disorder*.

**Figure 3.**
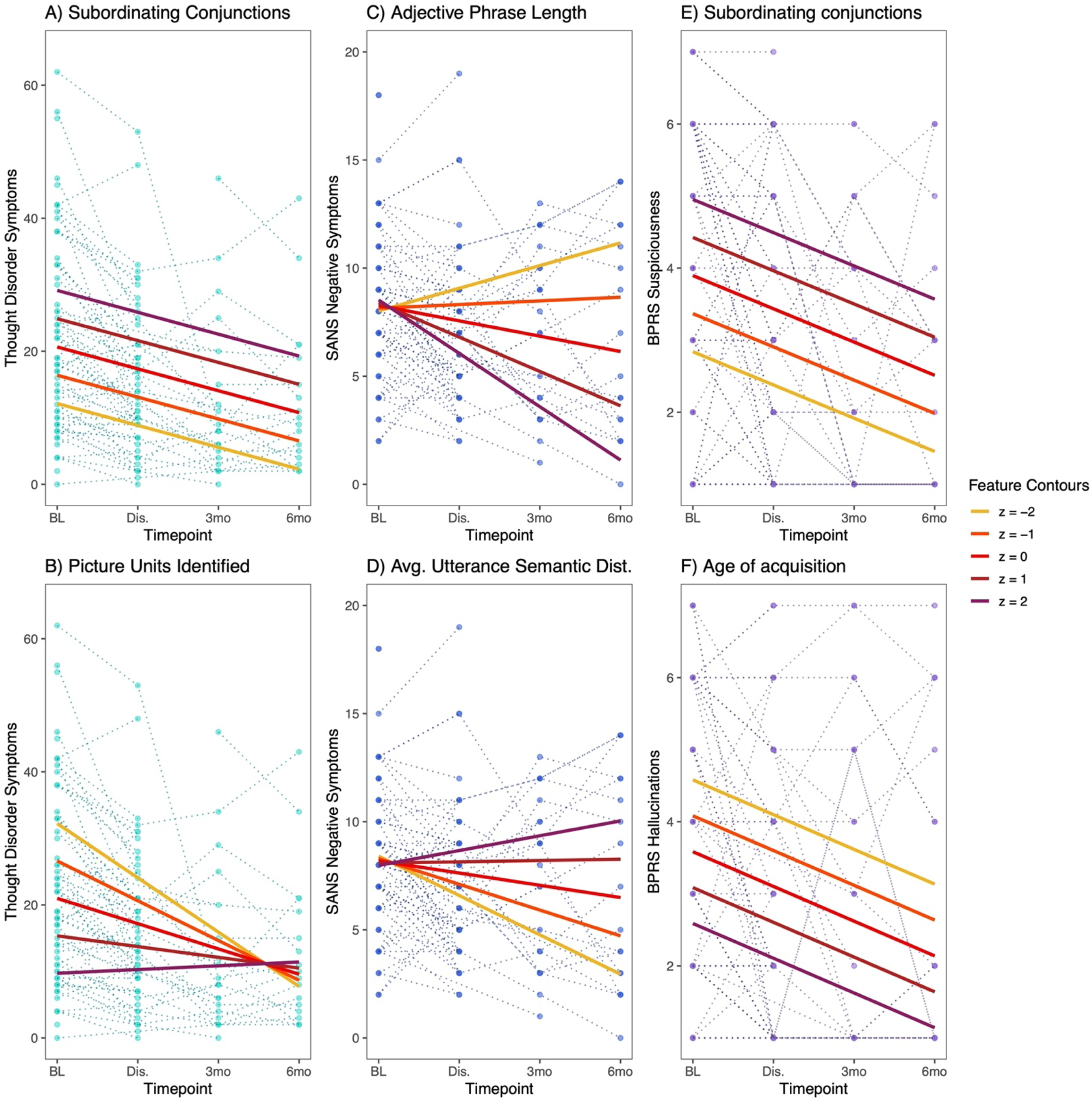
Psychosis Symptoms and Individual Speech and Language Features. Illustrative examples are shown for relationships between individual features and psychosis symptoms. Individual observations are plotted across the 4 timepoints (*Thought Disorder* – turquoise, *Negative Symptoms* – blue, *Positive Symptoms* – purple). Feature contours illustrate LMM predictions for each symptom domain at different values of the feature score (z=-2 to +2).

*Negative Symptoms* was significantly related to 14 features, all through linear interactions with timepoint. There was a general pattern of higher *Negative Symptoms* being related to features describing decreased speech complexity: words that are more easily visualized (imageability), less modifiers through adjectives and adverbs (e.g., Figure 3C), fewer connections (number of edges on speech graphs), and greater jumps in content (less elaboration, higher semantic distance between sentences; e.g., Figure 3D). As was true for the single component score, the individual features predicted greater variance in negative symptoms at follow-up.

None of the features were significantly related to *Positive Symptoms* after correction for multiple comparisons. Due to the heterogeneity of the domain, the individual items for *Suspiciousness, Hallucinations*, and *Unusual Thought Content* were examined. *Suspiciousness* was significantly related to 5 features, all through main effects. Higher *Suspiciousness* appears to be related to more subordinating conjunctions (e.g., Figure 3E) and less use of nouns in the journaling task. *Hallucinations* demonstrated 3 trend-level relationships which were included for illustrative purposes, including a relationship between using words with lower age of acquisition (learned at earlier age) and higher *Hallucinations* (Figure 3F). There were no significant relationships between speech and language features and *Unusual Thought Content*.

### Interactions with Gender and Race

Interactions between the speech and language parameter of interest and demographic variables were examined for the single component score and for the individual features highlighted as illustrative examples in Figure 3 (model structure detailed in Supplemental Table 2). There were no significant interactions between gender and any of the speech and language features (though there were main effects, they did not impact the relationship with speech features). For race, there were significant interactions with the single component score in *Negative Symptoms* and *Positive Symptoms*, and also with average utterance semantic distance in *Positive Symptoms* (Supplemental Table 4). The effect of race appeared to be primarily driven by a difference between White/Caucasian participants and all other groups, with highly divergent patterns (Supplemental Figure 1). There were no interactions for race and any of the features evaluated for *Thought Disorder, Suspiciousness*, or *Hallucinations*, nor for the other individual features predicting *Negative* and *Positive Symptoms*.

## Discussion

In this study, we first demonstrated that each of the major domains of psychosis symptoms was longitudinally related to speech and language features, on a global level. That is, taking a single component score that represented 357 features, we found that objective speech and language features were related to psychosis symptoms across four timepoints. This was true for *Thought Disorder, Negative Symptoms*, and *Positive Symptoms*. However, different patterns in the relationships could be observed – for example, with higher component scores being related to higher *Thought Disorder* but decreased *Negative Symptoms*. Next, we examined individual features to better understand how psychosis symptoms are related to specific measures obtained from speech and language, and found many significant relationships particularly for *Thought Disorder* and *Negative Symptoms*. Some features were related to multiple symptom areas (e.g., subordinating conjunctions, semantic coherence, imageability), though sometimes through different patterns, while other features were unique to one symptom domain. Taken together, it appears that each of the major psychosis symptom domains can be predicted by concurrently-measured speech and language features as a whole. It also seems promising that different kinds of speech and language measures can be combined to provide specificity for different kinds of psychosis symptoms. In fact, individual positive symptoms may be better predicted by speech features than the more heterogeneous construct of the *Positive Symptoms* domain.

When examining individual speech and language features, we found potentially interpretable patterns in their relationships with the psychosis symptom domains. *Thought Disorder* was related to several features reflecting more subordinated constructions (structures adding on additional information to the ongoing sentence) and less efficient identification of entities in picture descriptions. Though further work is needed to fully understand the connections between behavior and automated features, this might be interpreted as reflecting speech where ideas are (excessively) layered on top of one another, failing to communicate purposeful content. *Thought Disorder* was also related to decreased minimum distances in the semantic content of adjacent sentences, which would support the proposal of a ‘shrinking semantic space’ with shorter distances between concepts (25). *Negative Symptoms* on the other hand were related to several features describing a reduction in complexity on a word-choice level (higher imageability), syntactic level (fewer adjectives and adverbs), and discourse organization level (fewer edges connecting speech graphs). *Negative Symptoms* were additionally related to higher average semantic distance between adjacent sentences, particularly during the picture description task, which may reflect less elaboration and moving on to the next picture element instead of providing additional detail. Altogether, this fits with earlier work that links psychosis with decreased “semantic density” (26) and “idea density” (27).

The effect of timepoint on the relationships between features and symptoms is worth noting. Many of the highlighted models included a linear interaction between timepoint and feature, implying that the information provided by the speech and language features had different implications for symptom severity depending on the timing of the assessment – whether in a hospitalized acute setting, or after stabilization and discharge. This pattern is clearest for *Negative Symptoms*, where the single component score and all of the individual features demonstrated a pattern where very little variance is predicted during the first 2 timepoints, and greater variance is predicted during follow-up. A plausible explanation is that *Negative Symptoms* may be masked by more prominent *Positive Symptoms* and disorganization during acute psychosis exacerbations, as well as being superimposed upon sedation and medication side effects in the hospitalization setting. For *Thought Disorder* and *Positive Symptoms*, the single component score was described by a linear interaction with timepoint, but the majority of individual comparisons showed a main effect of the speech and language feature – i.e., variation in the speech feature had a consistent effect on the predicted symptom severity across timepoints, superimposed on an overall expectation of declining symptoms. Overall, these results suggest that phases of illness may affect some relationships between speech and psychosis symptoms, while others remain consistent. To our knowledge, this has not been previously examined, as most previous studies have focused on cross-sectional relationships.

In most cases, the relationships between psychosis symptoms and speech and language features were robust to the effects of gender and race. However, there were exceptions for race. It is unclear whether these were reproducible effects, or if these findings are driven by the relatively small sample of White/Caucasian participants. We are unaware of such demographic interactions having been previously tested or reported.

Many questions remain unanswered by the present study. While this is the largest longitudinal study of computational speech and language features and psychosis, to our knowledge, this study was not adequately powered to address some important concerns. We experienced a decline in participation for the later timepoints especially, which we attribute to the disruptive nature of an acute hospitalization event and pandemic-related considerations during the data collection period. Importantly, we focused on concurrent prediction of psychosis symptom severity with the speech and language features and did not predict outcomes in a prospective manner. The results also prompt us to question the appropriate granularity at which clinical constructs should be investigated. While the improved prediction of individual *Positive Symptoms* over the factor score would suggest higher accuracy with more detailed clinical targets, an over-specification may also increase sensitivity to assessment environment and individual participant variability. Perhaps different approaches will be optimal for different clinical applications.

The clinical implications are substantial for developing a scalable, cost-effective, low patient burden method of obtaining objective measurements of psychosis severity. Rehospitalizations are a major driver of poor outcomes in psychosis (17). A sensitive, efficient tool can be used to monitor patients for exacerbations between visits, and alert clinicians to intervene in a timely manner. Medication adherence is similarly critical in psychosis management, with side effects being a common reason for discontinuation (28). Speech and language biomarkers could potentially be used for more accurate and faster titration to the optimal dose and medication type, thereby decreasing patient distress, minimizing side effects, and improving adherence. Differential diagnosis also remains a challenge in some community and primary care settings (29), and could be aided by an objective biomarker to guide decision-making. Critically, due to the difficulty of demonstrating effectiveness for novel psychotherapeutics, there has been a slowing down of pharmaceutical investment in psychiatric disorders (30). Speech and language markers of psychosis severity can serve as objective outcome measures and facilitate the discovery and approval of novel effective pharmacologic and psychosocial treatments.

In total, our findings support the use of automated speech and language features as objective predictors for tracking psychosis symptoms severity. Different types of psychosis symptoms appear to be distinguishable with different speech and language measures. The present study is a critical initial step in deploying speech biomarkers for psychosis in a longitudinal context.

## Data Availability

Deidentified, processed data produced in the present study are available upon reasonable request to the authors.

## Acknowledgements

Funding for this study was received from Winterlight Labs, NIH K23 MH130750 (SXT), and the Brain and Behavior Research Foundation Young Investigator Grant (SXT). We thank the participants for their contributions, as well as: Anna Costakis, M.D., M.B.A., Hanna Contreras, B.A., Grace Serpe, B.A., Laura Chung, M.D., Mengdan Xu, M.Sc., Danielle De Souza, Ph.D., Jessica Robin, Ph.D., Yan Cong, Ph.D., Sunghye Cho, Ph.D., Sameer Pradhan, Ph.D., Aarush Mehta, and Amir Nikzad, M.D.

## References

1. Perålå J, Suvisaari J, Saarni SI, Kuoppasalmi K, Isometså E, Pirkola S, et al. Lifetime Prevalence of Psychotic and Bipolar I Disorders in a General Population. Arch Gen Psychiatry. 2007 Jan 1;64(1):19.

2. Tandon R, Keshavan MS, Nasrallah HA. Schizophrenia, “Just the Facts”: What we know in 2008. Part 1: Overview. Schizophrenia Research. 2008;100(1–3):4–19.

3. Zhang JP, Gallego JA, Robinson DG, Malhotra AK, Kane JM, Correll CU. Efficacy and safety of individual second-generation vs. first-generation antipsychotics in first-episode psychosis: a systematic review and meta-analysis. International Journal of Neuropsychopharmacology. 2013 Jul 1;16(6):1205–18.

4. Mueser KT, Deavers F, Penn DL, Cassisi JE. Psychosocial treatments for schizophrenia. Annual Review of Clinical Psychology. 2013;9(February):465–97.

5. Robinson DG, Schooler NR, Marcy P, Gibbons RD, Hendricks Brown C, John M, et al. Outcomes During and After Early Intervention Services for First-Episode Psychosis: Results Over 5 Years From the RAISE-ETP Site-Randomized Trial. Schizophrenia Bulletin. 2022 Sep 1;48(5):1021–31.

6. Ozomaro U, Wahlestedt C, Nemeroff CB. Personalized medicine in psychiatry: Problems and promises. BMC Medicine [Internet]. 2013 May 16 [cited 2020 Oct 26];11(1). Available from: https://pubmed-ncbi-nlm-nih-gov.proxy.library.upenn.edu/23680237/

7. Corcoran CM, Mittal VA, Bearden CE E. Gur R, Hitczenko K, Bilgrami Z, et al. Language as a biomarker for psychosis: A natural language processing approach. Schizophrenia Research. 2020;226:158–66.

8. Tang SX, Kriz R, Cho S, Park SJ, Harowitz J, Gur RE, et al. Natural Language Processing Methods are Sensitive to Sub-Clinical Linguistic Differences in Schizophrenia Spectrum Disorders. npj Schizophrenia. 2021;7:25.

9. Corcoran CM, Carrillo F, Fernández-Slezak D, Bedi G, Klim C, Javitt DC, et al. Prediction of psychosis across protocols and risk cohorts using automated language analysis. World Psychiatry. 2018;17(1):67–75.

10. Tang SX, Hånsel K, Cong Y, Nikzad AH, Mehta A, Cho S, et al. Latent Factors of Language Disturbance and Relationships to Quantitative Speech Features. Schizophrenia Bulletin. 2023 Mar 22;49(Supplement_2):S93–103.

11. Tang SX, Cong Y, Nikzad AH, Mehta A, Cho S, Hånsel K, et al. Clinical and computational speech measures are associated with social cognition in schizophrenia spectrum disorders. Schizophrenia Research. 2022;2023 Sep(259):28–37.

12. Krell R, Tang W, Hånsel K, Sobolev M, Cho S, Tang SX. Lexical and Acoustic Correlates of Clinical Speech Disturbance in Schizophrenia. W3PHAI 2021 Studies in Computational Intelligence. 2021;1013:9.

13. Nikzad AH, Cong Y, Berretta S, Hånsel K, Cho S, Pradhan S, et al. Who does what to whom? graph representations of action-predication in speech relate to psychopathological dimensions of psychosis. Schizophr. 2022 Dec;8(1):58.

14. Girard JM, Vail AK, Liebenthal E, Brown K, Kilciksiz CM, Pennant L, et al. Computational analysis of spoken language in acute psychosis and mania. Schizophrenia Research. 2022 Jul;245:97–115.

15. Goldberg TE, Dodge M, Aloia M, Egan MF, Weinberger DR. Effects of neuroleptic medications on speech disorganization in schizophrenia: biasing associative networks towards meaning. Psychol Med. 2000 Sep;30(5):1123–30.

16. Wang D, Gopal S, Baker S, Narayan VA. Trajectories and changes in individual items of positive and negative syndrome scale among schizophrenia patients prior to impending relapse. npj Schizophr. 2018 Dec;4(1):10.

17. Robinson DG, Schooler NR, Rosenheck RA, Lin H, Sint KJ, Marcy P, et al. Predictors of Hospitalization of Individuals With First-Episode Psychosis: Data From a 2-Year Follow-Up of the RAISE-ETP. PS. 2019 Jul;70(7):569–77.

18. First MB, Gibbon M. The structured clinical interview for DSM-IV axis I disorders (SCID-I) and the structured clinical interview for DSM-IV axis II disorders (SCID-II). Comprehensive handbook of psychological assessment. 2004;2:134–43.

19. Andreasen NC. Scale for the Assessment of Thought, Language, and Communication (TLC). Schizophrenia Bulletin. 1986 Jan 1;12(3):473–82.

20. Andreasen NC. Scale for the Assessment of Negative Symptoms (SANS). In Iowa City: University of Iowa; 1984.

21. Robinson D, Woerner M, Schooler N. Intervention Research in Psychosis: Issues Related to Clinical Assessment. Schizophrenia Bulletin. 2000 Jan 1;26(3):551–6.

22. Overall JE, Hollister L, Pichot P. Major Psychiatric Disorders: A Four-Dimensional Model. Arch Gen Psychiatry. 1967 Feb 1;16(2):146.

23. Pinheiro J, Bates D, DebRoy S, Sarkar D, R Core Team. nlme: Linear and Nonlinear Mixed Effects Models [Internet]. 2022. Available from: https://CRAN.R-project.org/package=nlme

24. Benjamini Y, Hochberg Y. Controlling the False Discovery Rate: A Practical and Powerful Approach to Multiple Testing. Journal of the Royal Statistical Society: Series B. 1995;57(1):289–300.

25. He R, Palominos C, Zhang H, Alonso-Sánchez MF, Palaniyappan L, Hinzen W. Navigating the semantic space: Unraveling the structure of meaning in psychosis using different computational language models. Psychiatry Research. 2024 Mar;333:115752.

26. Rezaii N, Walker E, Wolff P. A machine learning approach to predicting psychosis using semantic density and latent content analysis. npj Schizophrenia. 2019;5(1):9.

27. Moe AM, Breitborde NJK, Shakeel MK, Gallagher CJ, Docherty NM. Idea density in the life-stories of people with schizophrenia: Associations with narrative qualities and psychiatric symptoms. Schizophrenia Research. 2016;172(1–3):201–5.

28. Robinson DG, Woerner MG, Alvir JMaJ, Bilder RM, Hinrichsen GA, Lieberman JA. Predictors of medication discontinuation by patients with first-episode schizophrenia and schizoaffective disorder. Schizophrenia Research. 2002 Oct;57(2–3):209–19.

29. Al-Huthail YR. Accuracy of Referring Psychiatric Diagnosis. Int J Health Sci (Qassim). 2008 Jan;2(1):35–8.

30. O’Brien PL, Thomas CP, Hodgkin D, Levit K, Mark TL. The diminished pipeline for medications to treat mental health and substance use disorders. Psychiatr Serv. 2014 Dec 1;65(12):1433–8.

