## Supplemental Materials for "Automated, Objective Speech and Language Markers of Longitudinal Changes in Psychosis Symptoms"

Sunny X. Tang, M.D., Michael J. Spilka, Ph.D., Majnu John, Ph.D., Michael L. Birnbaum, M.D., Ema Saito, M.D., Sarah A. Berretta, B.A., Leily M. Behbehani, B.S., Mark Y. Liberman, Ph.D., Anil K. Malhotra, M.D., William Simpson, Ph.D., John M. Kane, M.D.

***Contents*** ***Page***

#### Supplemental Table 1 – Top Loading Features from PCA (n=50)

| Feature Name | Description | Example | Loading |
|---|---|---|---|
| Personal pronoun noun phrase (PIC) | Noun phrase productions consisting only of a personal pronoun, expressed as a fraction of all syntactic productions (picture description) | [NP [PRP I]] am going to wear a mask. | 0.67 |
| Subordinate clause: preposition + sentence (PIC) | Subordinating clause productions consisting of a preposition and sentence, expressed as a fraction of all syntactic productions (picture description) | [SBAR [IN in] [S the house with the chimney]] | 0.57 |
| Subordinate clause: preposition + sentence (JOU) | Subordinating clause productions consisting of a preposition and sentence, expressed as a fraction of all syntactic productions (journaling) | [SBAR [IN in] [S the house with the chimney]] | 0.55 |
| Subordinating conjunctions (JOU) | Fraction of words that are subordinating conjunctions (journaling) | "because", "although" | 0.54 |
| Dependent clauses proportion (JOU) | Number of dependent clauses per clause (journaling) | [C I saw someone today] [DC with whom I used to play soccer]. [C It was such a surprise]! (1/3) | 0.53 |
| Subordinating conjunctions count (PIC) | Number of words labelled as subordinating conjunctions using part-of-speech tagging, expressed as a fraction of total words (picture description) | "because", "although" | 0.52 |
| Age of acquisition (JOU) | Word age of acquisition (according to normative data), averaged over all words in the transcript | NA | 0.5 |
| Total words (JOU) | Fraction of annotations that are words, out of all annotations including pauses and non-word utterances (journaling) | NA | 0.49 |
| Verb phrase: modal verb and verb phrase (PIC) | Verb phrase productions consisting of a modal and a verb phrase, expressed as a fraction of all syntactic productions (picture description) | You [VP [M can] [VP take what you need]] | 0.49 |
| Propositional density ratio (PIC) | Fraction of words that are elementary propositions (including verbs, adverbs, adjectives, conjunctions, and prepositions) multiplied by 10 (picture description) | The quick(1) brown(2) fox jumped(3) over the lazy(4) dog because(5) she was feeling(6) ambitious(7) | 0.49 |
| Adverbial phrase proportion (PIC) | Proportion of words belonging to an adverbial phrase (picture description) | [S[ADVP By the time we get home] [NP the sun] [VP will have gone down]] (6) | 0.48 |
| Age of acquisition: nouns (JOU) | Word age of acquisition (according to normative data), averaged over all nouns in the transcript (journaling) | NA | 0.47 |
| Function words (PIC) | Fraction of words that are function words, including conjunctions, determiners, prepositions, modal verbs, particles, pronouns (picture description) | "and", "if", "but", "when", "because" | 0.46 |

#### Supplemental Table 1 – Top-Loading Features from PCA (Continued)

| Feature Name | Description | Example | Loading |
|---|---|---|---|
| Average adverbial phrase length (PIC) | Average length of adverbial phrases (picture description) | [S[ADVP By the time we get home] [NP the sun] [VP will have gone down]] (6) | 0.45 |
| Adverbs (JOU) | Number of words labelled as adverbs using part-of-speech tagging, expressed as a fraction of total words (journaling) | "quickly", "hurriedly" | 0.45 |
| Prepositional phrase length (JOU) | Average length of prepositional phrases (journaling) | [PP underneath the bright lights] (4) | 0.44 |
| Prepositional phrase length (PIC) | Average length of prepositional phrases (picture description) | [PP underneath the bright lights] (4) | 0.44 |
| Adjective phrase length (PIC) | Average length of adjectival phrases (picture description) | [ADJP brewed by monks] (3) | 0.44 |
| Subordinating conjunctions count (JOU) | Number of words labelled as subordinating conjunctions using part-of-speech tagging, expressed as a fraction of total words (journaling) | "because", "although" | 0.44 |
| Verb phrase: gerund verb + subordinate or relative clause (PIC) | Verb phrase productions consisting of an inflected verb and a subordinate or relative clause (picture description) | I [VP [VBP hope] [SBAR that the cat will sleep]] | 0.43 |
| Age of acquisition (PIC) | Word age of acquisition (according to normative data) averaged over all words in the transcript (picture description) | NA | 0.43 |
| Verb phrase: modal verb and verb phrase (PIC) | Verb phrase productions consisting of a verb in base form (infinitive) and a subordinate or relative clause, expressed as a fraction of all syntactic productions (picture description) | I came [VP [VB to dance] [SBAR even though there is no music]] | 0.42 |
| Subordinating conjunctions (PIC) | Fraction of words that are subordinating conjunctions (picture description) | "because", "although" | 0.41 |
| Frequency (PIC) | Word frequency of usage (according to normative data), averaged over all words in the transcript (picture description) | NA | 0.41 |
| Demonstratives (PIC) | Fraction of words that are demonstratives (picture description) | "this", "that", "these", "those" | 0.41 |
| Subordinate clause: sentence (PIC) | Relative or subordinate clause productions that are standalone sentences, expressed as a fraction of all syntactic productions (picture description) | [while listeninig to music] | 0.41 |
| Subordinate to coordinate ratio (JOU) | Ratio of subordinate to coordinate conjunctions (journaling) | NA | 0.4 |
| Noun phrase: Noun phrase + clause (JOU) | Noun phrases consisting of noun phrase and an embedded clause, relative clause, or opener clause; expressed as a fraction of all syntactic productions (journaling) | This is [NP [NP the one ] [SBAR that I was talking about]] | 0.4 |

#### Supplemental Table 1 – Top-Loading Features from PCA (Continued)

| Feature Name | Description | Example | Loading |
|---|---|---|---|
| Total audio duration (JOU) | Total duration of the audio recording (journaling) | NA | 0.39 |
| Verb phrase: gerund verb + noun phrase (JOU) | Verb phrases consisting of gerund verb and noun phrase, expressed as a fraction of all syntactic productions (journaling) | I am [VP [VBG drinking] [NP your hot chocolate]] | -0.39 |
| Verb imageability (JOU) | Word imageability score (according to normative data), averaged over all verbs in the transcript (journaling) | NA | -0.4 |
| Noun phrase rate (JOU) | Number of noun phrases, expressed as a fraction of total words (journaling) | [NP The great tree] was located in [NP the middle] of [NP the forest] (7/11) | -0.4 |
| Noun phrase: Noun (JOU) | Noun phrases consisting only of a singular noun, expressed as a fraction of all syntactic productions (journaling) | [NP[NN Winter] | -0.4 |
| Picture units identified:Action (PIC) | Fraction of all words that are correctly mentioned actions represented in the stimulus picture (picture description) | NA | -0.4 |
| Number of edges (JOU) | Number of edges (connections between successive content words) in the speech graph divided by the total number of words (journaling) | NA | -0.41 |
| Honore (PIC) | A measure of type-to-token ratio (vocabulary richness) that accounts for words used only once (picture description) | NA | -0.41 |
| Imageability (PIC) | Word imageability score (according to normative data), averaged over all words in the transcript (picture description) | NA | -0.44 |
| Noun imageability (JOU) | Word imageability score (according to normative data), averaged over all nouns in the transcript (journaling) | NA | -0.46 |
| Imageability (JOU) | Word imageability score (according to normative data), averaged over all words in the transcript (journaling) | NA | -0.47 |
| Minimum cosine distance (PIC) | Minimum cosine distance between pairs of utterence vector representations (the lower this value, the more similar are the most similar pair of utterances) (picture description) | NA | -0.48 |
| Pause to word ratio (JOU) | Number of pauses divided by the number of words (journaling) | NA | -0.48 |
| Number of sentence productions (PIC) | Number of sentence productions, expressed as a fraction of all syntactic productions (picture description) | [S I am doing a journaling task] | -0.49 |
| Minimum local coherence distance (fastText 300) (JOU) | Minimum cosine distance between successive utterances using the fastText word vector representation model with 300 dimensions (journaling) | NA | -0.54 |
| Unfilled pauses (JOU) | Unfilled pauses (silences), expressed as a fraction of all transcript annotations (journaling) | NA | -0.54 |

#### Supplemental Table 1 – Top-Loading Features from PCA (Continued)

| Feature Name | Description | Example | Loading |
|---|---|---|---|
| Number of sentence productions (JOU) | Number of sentence productions, expressed as a fraction of all syntactic productions (journaling) | [S I am doing a picture description task] | -0.57 |
| Minimum utterance semantic distance (fastText) (PIC) | Minimum cosine distance between successive utterances using the fastText word vector representation model with 300 dimensions (picture description) | NA | -0.61 |
| Minimum utterance semantic distance (Google) (JOU) | Minimum cosine distance between successive utterances using the Google word2vec word vector representation model with 300 dimensions (journaling) | NA | -0.61 |
| Average utterance semantic distance (Google) (PIC) | Average cosine distance between successive utterances using the Google word2vec word vector representation model with 300 dimensions (picture) | NA | -0.65 |
| Picture units identified: subject (PIC) | Fraction of all words that are correctly mentioned subjects represented in the stimulus picture (picture description) | NA | -0.65 |
| Picture units identified (PIC) | Fraction of all words that are correctly mentioned details represented in the stimulus picture (picture description) | NA | -0.69 |

| Abbreviation | Definition |
|---|---|
| ADJP | Adjective phrase |
| ADVP | Adverb phrase |
| IN | Preposition or subordinating conjunction |
| JOU | Journaling task |
| M | Modal verb |
| NP | Noun phrase |
| PIC | Picture Description task |
| PRP | Personal pronoun |
| S | Sentence |
| SBAR | Subordinate or relative clause |
| VB | Verb base form (infinitive) |
| VBG | Verb gerund |
| VBP | Verb non 3rd person singular present (inflected) |
| VP | Verb phrase |

#### Supplemental Table 2 – Linear Mixed Models (LMMs) Structures

| Model Type | Model Structure | Parameter of Interest |
|---|---|---|
| **Main Effect** | Outcome ~ Timepoint + Feature + 1\|Participant | Feature |
| **Linear Interaction** | Outcome ~ Timepoint + Feature + Timepoint*Feature + 1\|Participant | Timepoint*Feature |
| **Quadratic Interaction** | Outcome ~ Timepoint + Timepoint^2 + Feature + Timepoint*Feature + Timepoint^2*Feature + 1\|Participant | Timepoint^2*Feature |
| **Demographic Interaction** | Outcome ~ Timepoint + Feature + DemographicVariable + Timepoint*Feature + Timepoint*DemographicVariable + Feature*DemographicVariable + Timepoint*Feature*DemographicVariable + 1\|Participant | Timepoint*Feature*DemographicVariable |

*Note*: All models were random intercept linear mixed models where random effects were by participant.

### Supplemental Table 3 – Unabbreviated LMM Results

##### A. Features Relating to Thought Disorder

| | | *Int.* | *Timept.* | *Timesq.* | *Main Effect* | | | *Lin. Interaction* | | | *Quad. Interaction* | | *Model* | |
|---|---|---|---|---|---|---|---|---|---|---|---|---|---|---|
| ***Feature*** | ***Model*** | ***Coeff.*** | ***Coeff.*** | ***Coeff.*** | ***Coeff.*** | ***p*** | ***p-adj.*** | ***Coeff.*** | ***p*** | ***p-adj.*** | ***Coeff.*** | ***p*** | ***AIC*** | ***BIC*** |
| Single component score | M | 20.3 | -2.9 | | 4.2 | <0.001 | | | | | | | 1220 | 1235 |
| Single component score | L | 20.2 | -3.4 | | 5.9 | <0.001 | | -2.1 | <0.001 | | | | 1212 | 1230 |
| Single component score | Q | 21.0 | -6.2 | 1.1 | 5.4 | <0.001 | | -1.0 | 0.61 | | -0.3 | 0.67 | 1210 | 1235 |
| Subordinating conjunctions (JOU) | M | 20.6 | -3.3 | | 4.2 | <0.001 | <0.001 | | | | | | 1212 | 1227 |
| Min. utterance semantic dist. (Google) (JOU) | M | 20.8 | -3.5 | | -3.5 | <0.001 | 0.001 | | | | | | 1216 | 1231 |
| Min. utterance semantic distance (fastText) (PIC) | M | 20.9 | -3.7 | | -3.3 | <0.001 | 0.002 | | | | | | 1218 | 1233 |
| Picture units identified:Action (PIC) | M | 21.6 | -4.3 | | -3.6 | <0.001 | 0.002 | | | | | | 1218 | 1234 |
| Total audio duration (JOU) | M | 20.8 | -3.5 | | 3.5 | <0.001 | 0.002 | | | | | | 1219 | 1234 |
| Subordinating conjunctions count (JOU) | M | 21.0 | -3.6 | | 2.9 | <0.001 | 0.004 | | | | | | 1221 | 1236 |
| Subordinate clause: preposition + sentence (JOU) | M | 21.1 | -3.6 | | 2.9 | 0.004 | 0.02 | | | | | | 1225 | 1240 |
| Subordinate clause: sentence (PIC) | M | 20.8 | -3.4 | | 2.6 | 0.005 | 0.03 | | | | | | 1225 | 1241 |
| Subordinating conjunctions (PIC) | M | 21.0 | -3.6 | | 2.4 | 0.006 | 0.03 | | | | | | 1226 | 1241 |
| Imageability (JOU) | M | 20.7 | -3.3 | | -2.6 | 0.006 | 0.03 | | | | | | 1226 | 1241 |
| Picture units identified (PIC) | L | 21.0 | -3.8 | | -5.6 | <0.001 | | 2.2 | <0.001 | 0.03 | | | 1214 | 1232 |
| Picture units identified: subject (PIC) | L | 21.5 | -4.2 | | -3.5 | 0.003 | | 2.3 | 0.001 | 0.03 | | | 1222 | 1240 |
| Noun phrase: Noun (JOU) | L | 21.1 | -4 | | -4.3 | <0.001 | | 2.3 | 0.002 | 0.03 | | | 1219 | 1238 |
| Dependent clauses proportion (JOU) | M | 21.2 | -3.7 | | 2.1 | 0.01 | 0.05 | | | | | | 1227 | 1242 |
| Subordinate to coordinate ratio (JOU) | M | 21.1 | -3.5 | | 2.2 | 0.01 | 0.05 | | | | | | 1227 | 1242 |

#### Supplemental Table 3 – Unabbreviated LMM Results (continued)

**B. Features Relating to Negative Symptoms**

| *Feature* | *Model* | *Int. Coeff.* | *Timept. Coeff.* | *Timesq. Coeff.* | *Main Effect Coeff.* | *Main Effect p* | *Main Effect p-adj.* | *Lin. Interaction Coeff.* | *Lin. Interaction p* | *Lin. Interaction p-adj.* | *Quad. Interaction Coeff.* | *Quad. Interaction p* | *Model AIC* | *Model BIC* |
|---|---|---|---|---|---|---|---|---|---|---|---|---|---|---|
| Single component score | M | 8.3 | -0.6 | | -0.6 | 0.08 | | | | | | | 857 | 872 |
| Single component score | L | 8.2 | -0.8 | | 0.3 | 0.38 | | -1.0 | <0.001 | | | | 844 | 862 |
| Single component score | Q | 8.2 | -0.6 | -0.1 | 0.3 | 0.42 | | -1.0 | 0.14 | | 0.0 | 0.99 | 850 | 874 |
| Subordinate clause: preposition + sentence (JOU) | L | 8.2 | -0.6 | | 1.1 | 0.003 | | -0.9 | <0.001 | 0.02 | | | 850 | 868 |
| Min. utterance semantic distance (fastText) (PIC) | L | 8.2 | -0.6 | | -0.4 | 0.2 | | 0.7 | 0.001 | 0.02 | | | 851 | 870 |
| Imageability (PIC) | L | 8.3 | -0.5 | | -0.5 | 0.2 | | 0.7 | 0.002 | 0.02 | | | 852 | 870 |
| Age of acquisition (PIC) | L | 8.1 | -0.4 | | 0.6 | 0.1 | | -0.7 | 0.002 | 0.02 | | | 853 | 871 |
| Adjective phrase length (PIC) | L | 8.3 | -0.7 | | 0.1 | 0.7 | | -0.9 | 0.003 | 0.02 | | | 847 | 865 |
| Number of sentence productions (JOU) | L | 8.1 | -0.6 | | -0.5 | 0.2 | | 0.8 | 0.003 | 0.02 | | | 853 | 871 |
| Adverbs (JOU) | L | 8.1 | -0.3 | | 0.4 | 0.2 | | -0.8 | 0.003 | 0.02 | | | 851 | 870 |
| Average utterance semantic distance (Google) (PIC) | L | 8.2 | -0.6 | | -0.1 | 0.8 | | 0.6 | 0.003 | 0.02 | | | 850 | 868 |
| Number of edges (JOU) | L | 8.1 | -0.5 | | -0.7 | 0.05 | | 0.9 | 0.004 | 0.02 | | | 854 | 872 |
| Subordinating conjunctions (JOU) | L | 8.1 | -0.6 | | 1 | 0.006 | | -0.8 | 0.004 | 0.02 | | | 853 | 872 |
| Min. utterance semantic dist. (Google) (JOU) | L | 8.2 | -0.6 | | -0.5 | 0.1 | | 0.7 | 0.009 | 0.04 | | | 856 | 874 |
| Noun phrase: Noun phrase + clause (JOU) | L | 8.1 | -0.5 | | 0.6 | 0.08 | | -0.6 | 0.01 | 0.04 | | | 856 | 874 |
| Verb phrase: modal verb and verb phrase (PIC) | L | 8.2 | -0.6 | | 0.1 | 0.8 | | -0.7 | 0.01 | 0.04 | | | 854 | 872 |
| Function words (PIC) | L | 8.2 | -0.5 | | 0.4 | 0.3 | | -0.5 | 0.01 | 0.05 | | | 856 | 874 |

#### Supplemental Table 3 – Unabbreviated LMM Results (continued)

##### C. Features Relating to Positive Symptoms

| Feature | Model | Int. Coeff. | Timept. Coeff. | Timesq. Coeff. | Main Effect Coeff. | Main Effect p | Main Effect p-adj. | Lin. Interaction Coeff. | Lin. Interaction p | Lin. Interaction p-adj. | Quad. Interaction Coeff. | Quad. Interaction p | Model AIC | Model BIC |
|---|---|---|---|---|---|---|---|---|---|---|---|---|---|---|
| Single component score | M | 20.2 | -2.4 | | 0.7 | 0.20 | | | | | | | 999 | 1015 |
| Single component score | L | 20.2 | -2.6 | | 1.3 | 0.03 | | -0.8 | 0.04 | | | | 997 | 1015 |
| Single component score | Q | 20.6 | -3.9 | 0.5 | 0.8 | 0.20 | | 0.6 | 0.54 | | -0.5 | 0.20 | 998 | 1022 |

##### D. Features Relating to Suspiciousness

| Feature | Model | Int. Coeff. | Timept. Coeff. | Timesq. Coeff. | Main Effect Coeff. | Main Effect p | Main Effect p-adj. | Lin. Interaction Coeff. | Lin. Interaction p | Lin. Interaction p-adj. | Quad. Interaction Coeff. | Quad. Interaction p | Model AIC | Model BIC |
|---|---|---|---|---|---|---|---|---|---|---|---|---|---|---|
| Subordinating conjunctions (JOU) | M | 3.9 | -0.5 | | 0.5 | 0.001 | 0.03 | | | | | | 663 | 678 |
| Noun phrase rate (JOU) | M | 3.9 | -0.5 | | -0.5 | 0.002 | 0.03 | | | | | | 663 | 678 |
| Noun phrase: Noun (JOU) | M | 3.9 | -0.5 | | -0.5 | 0.002 | 0.03 | | | | | | 663 | 678 |
| Imageability (JOU) | M | 3.9 | -0.4 | | -0.5 | 0.003 | 0.04 | | | | | | 664 | 680 |
| Number of edges (JOU) | M | 3.9 | -0.5 | | -0.4 | 0.005 | 0.05 | | | | | | 665 | 680 |

##### E. Features Relating to Hallucinations

| Feature | Model | Int. Coeff. | Timept. Coeff. | Timesq. Coeff. | Main Effect Coeff. | Main Effect p | Main Effect p-adj. | Lin. Interaction Coeff. | Lin. Interaction p | Lin. Interaction p-adj. | Quad. Interaction Coeff. | Quad. Interaction p | Model AIC | Model BIC |
|---|---|---|---|---|---|---|---|---|---|---|---|---|---|---|
| Age of acquisition: nouns (JOU) | M | 3.6 | -0.5 | | -0.5 | 0.001 | 0.06 | | | | | | 645 | 660 |
| Min. utterance semantic distance (fastText) (PIC) | M | 3.5 | -0.5 | | 0.5 | 0.003 | 0.06 | | | | | | 646 | 662 |
| Average utterance semantic distance (Google) (PIC) | M | 3.5 | -0.4 | | 0.5 | 0.003 | 0.06 | | | | | | 646 | 662 |

*Note:* M – main effect; L – linear effect; Q – quadratic effect; Coeff. – coefficient; JOU – Journaling task; PIC – Picture description task; AIC - Akaike Information Criteria; BIC – Bayesian Information Criteria.

#### Supplemental Table 4 – Interactions with Gender and Race

**A. Thought Disorder Demographic Interactions**

| *Demo Variable* | *Feature* | *Interaction Term* | *Coeff.* | *p* | *AIC* | *BIC* |
|---|---|---|---|---|---|---|
| Gender | Component Score | Woman*Timepoint*ComponentScore | -3.6 | 0.35 | 1212 | 1230 |
| | | Asian*Timepoint*ComponentScore | 0.0 | 1.00 | | |
| Race | Component Score | OtherRace*Timepoint*ComponentScore | -1.3 | 0.85 | 1162 | 1215 |
| | | White*Timepoint*ComponentScore | 2.3 | 0.29 | | |
| Gender | Subordinating conjunctions (JOU) | Woman*Feature | 1.1 | 0.62 | 1090 | 1110 |
| | | Asian*Feature | -2.3 | 0.33 | | |
| Race | Subordinating conjunctions (JOU) | OtherRace*Feature | -4.7 | 0.12 | 1179 | 1212 |
| | | White*Feature | -3.4 | 0.15 | | |
| Gender | Picture units identified:Action (PIC) | Woman*Timepoint*Feature | 3.2 | 0.28 | 1088 | 1118 |
| | | Asian*Timepoint*Feature | 1.0 | 0.62 | | |
| Race | Picture units identified:Action (PIC) | OtherRace*Timepoint*Feature | 1.0 | 0.83 | 1166 | 1219 |
| | | White*Timepoint*Feature | 1.9 | 0.33 | | |

**B. Negative Symptoms Demographic Interactions**

| *Demo Variable* | *Feature* | *Interaction Term* | *Coeff.* | *p* | *AIC* | *BIC* |
|---|---|---|---|---|---|---|
| Gender | Component Score | Woman*Timepoint*ComponentScore | 0.2 | 0.88 | 779 | 808 |
| | | Asian*Timepoint*ComponentScore | -0.1 | 0.84 | | |
| Race | Component Score | OtherRace*Timepoint*ComponentScore | -0.4 | 0.86 | 827 | 881 |
| | | White*Timepoint*ComponentScore | 1.7 | 0.02 | | |
| White/Caucasian | Component Score | White*Timepoint*ComponentScore | 1.7 | 0.02 | 840 | 870 |
| Gender | Adjective phrase length (PIC) | Woman*Timepoint*Feature | -0.5 | 0.71 | 781 | 811 |
| | | Asian*Timepoint*Feature | 0.8 | 0.31 | | |
| Race | Adjective phrase length (PIC) | OtherRace*Timepoint*Feature | -1.3 | 0.58 | 833 | 886 |
| | | White*Timepoint*Feature | 1.2 | 0.24 | | |
| Gender | Average utterance semantic distance (Google) (PIC) | Woman*Timepoint*Feature | 0.6 | 0.69 | 784 | 814 |
| | | Asian*Timepoint*Feature | 0.9 | 0.14 | | |
| Race | Average utterance semantic distance (Google) (PIC) | OtherRace*Timepoint*Feature | 1.3 | 0.58 | 830 | 883 |
| | | White*Timepoint*Feature | -1.8 | 0.03 | | |

#### Supplemental Table 4 – Interactions with Gender and Race

**C. Positive Symptoms Demographic Interactions**

| *Demo Variable* | *Feature* | *Interaction Term* | *Coeff.* | *p* | *AIC* | *BIC* |
|---|---|---|---|---|---|---|
| Gender | Component Score | Woman*Timepoint*ComponentScore | 2.6 | 0.22 | 905 | 934 |
| | | Asian*Timepoint*ComponentScore | 0.1 | 0.88 | | |
| Race | Component Score | OtherRace*Timepoint*ComponentScore | -2.0 | 0.56 | 954 | 1008 |
| | | White*Timepoint*ComponentScore | 3.4 | 0.003 | | |
| White/Caucasian | Component Score | White*Timepoint*ComponentScore | 3.3 | 0.003 | 981 | 1011 |

**D. Suspiciousness Demographic Interactions**

| *Demo Variable* | *Feature* | *Interaction Term* | *Coeff.* | *p* | *AIC* | *BIC* |
|---|---|---|---|---|---|---|
| Gender | Subordinating conjunctions (JOU) | Woman*Feature | 0.7 | 0.09 | 611 | 632 |
| | | Asian*Feature | 0.2 | 0.65 | | |
| Race | Subordinating conjunctions (JOU) | OtherRace*Feature | 0.5 | 0.33 | 659 | 692 |
| | | White*Feature | -0.4 | 0.33 | | |

**E. Hallucinations Demographic Interactions**

| *Demo Variable* | *Feature* | *Interaction Term* | *Coeff.* | *p* | *AIC* | *BIC* |
|---|---|---|---|---|---|---|
| Gender | Age of acquisition: nouns (JOU) | Woman*Feature | 0.4 | 0.50 | 594 | 615 |
| | | Asian*Feature | 0.0 | 0.95 | | |
| Race | Age of acquisition: nouns (JOU) | OtherRace*Feature | 0.3 | 0.64 | 643 | 676 |
| | | White*Feature | 0.5 | 0.23 | | |

*Note:* Coeff. – coefficient; JOU – Journaling task; PIC – Picture description task; AIC -Akaike Information Criteria; BIC – Bayesian Information Criteria.

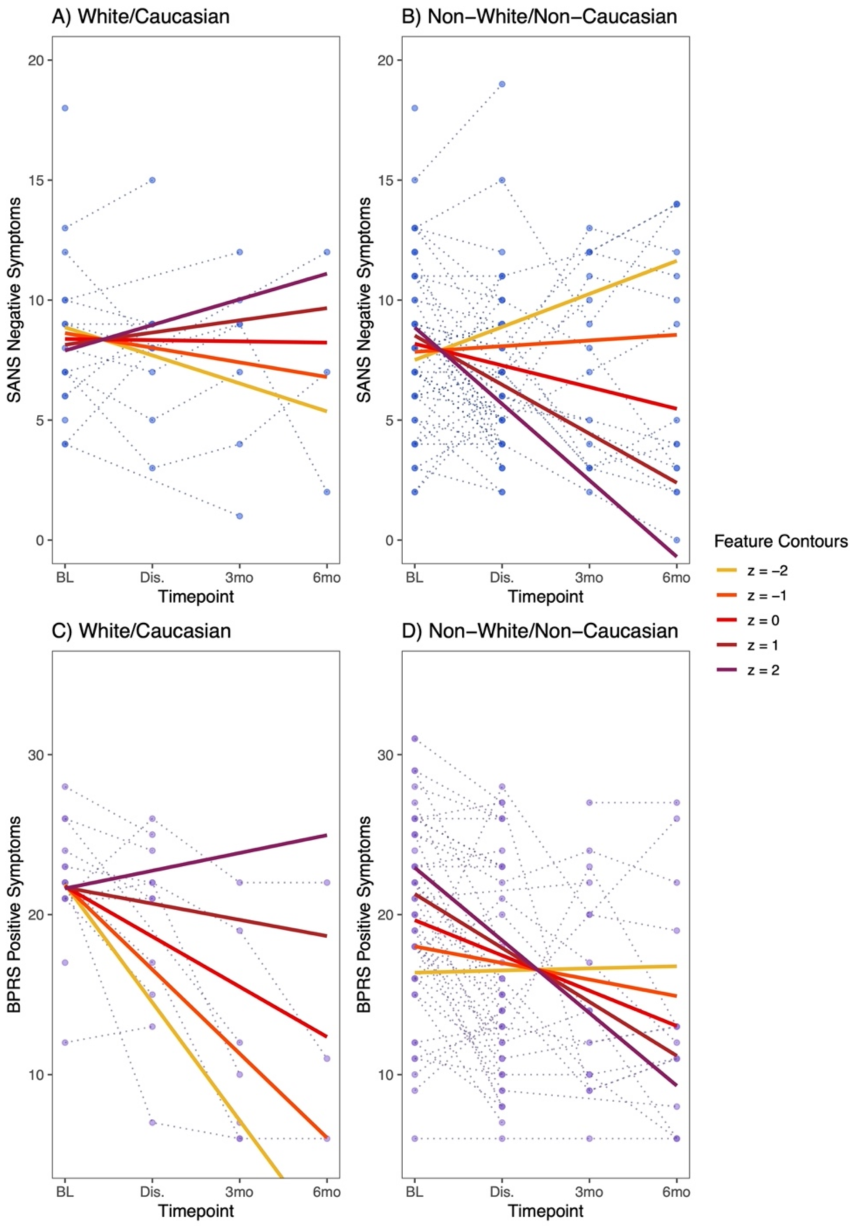

**Supplemental Figure 1 – Interactions between Single Component Score and Race.** Relationships between the single component score and *Negative Symptoms* are shown for A) White/Caucasian participants and B) Non-White/Non-Caucasian participants. Similarly, the relationships are shown for *Positive Symptoms* and C) White/Caucasian participants and D) Non-White/Non-Caucasian participants. Relationships between the component score and negative symptoms are reversed A) vs. B), and also highly divergent for C) vs. D). However, note the relative scarcity of data for White/Caucasian participants.
